# Comparative Effectiveness of CRT-P vs CRT-D in Octogenarians With HFrEF and LBBB: A Real-World, Propensity-Matched Cohort Study

**DOI:** 10.1101/2025.07.31.25332554

**Authors:** Festus Ibe, Justin Riley Lam, Phuuwadith Wattanachayakul, Emmanuel Otabor, Ikechukwu Ifedili, Kenechukwu Mezue, Irhoboudu Atogwe, Khraisha Ola, Behnam Bozorgnia

**Affiliations:** Department of Internal Medicine, Jefferson Einstein Hospital, Philadelphia, PA, USA; Sidney Kimmel Medical College, Thomas Jefferson University, Philadelphia, PA, USA; Department of Medicine, Division of Cardiovascular Disease, University of Tennessee Health Science Center, Memphis, TN, USA; Division of Cardiovascular Medicine, Yale School of Medicine, New Haven, CT, USA; Division of Cardiovascular disease, Jefferson Einstein Hospital, Philadelphia, PA, USA

**Keywords:** Cardiac Resynchronization Therapy, Octogenarians, Heart Failure with Reduced Ejection Fraction, Left Bundle Branch Block

## Abstract

**Background:** Cardiac resynchronization therapy (CRT) has demonstrated survival and symptom benefits in patients with heart failure with reduced ejection fraction (HFrEF). However, the comparative effectiveness of CRT with a defibrillator (CRT-D) versus pacemaker-only (CRT-P) remains uncertain in octogenarians, who are underrepresented in clinical trials.

**Objective:** To assess long-term mortality and hospitalization outcomes associated with CRT-D versus CRT-P in octogenarians with HFrEF and left bundle branch block (LBBB), using real-world propensity-matched data.

**Methods:** We identified 1,240 patients aged ≥80 years with HFrEF (LVEF ≤35%) and LBBB who underwent device implantation. After 1:1 propensity score matching across 65 clinical variables, 772 patients (386 CRT-D, 386 CRT-P) were included in the primary analysis. The primary endpoint was all-cause mortality over 3 years. Secondary endpoints included timepoint-specific mortality and all-cause hospitalization burden. Prespecified subgroup analyses were performed by ejection fraction (EF ≤25%, 26–35%) and by sex within each CRT modality.

**Results:** Three-year mortality did not differ significantly between CRT-D and CRT-P (37.2% vs 39.1%; HR 1.026, 95% CI 0.814–1.293; p = 0.829). Mortality at 30 days, 90 days, and 1 year was numerically lower in CRT-D but not statistically significant. CRT-D recipients experienced fewer hospitalizations (mean 4.05 vs 5.36; p = 0.032). In EF-stratified subgroups, mortality was modestly lower with CRT-D (risk difference –3.8% in EF ≤25%; –2.1% in EF 26–35%), though not significant. Among CRT-D recipients, females had lower mortality than males (20.0% vs 29.5%; HR 0.637, 95% CI 0.353–1.149).

**Conclusions:** In this real-world cohort of octogenarians with HFrEF, CRT-D did not improve survival over CRT-P but was associated with reduced hospitalization burden. These findings highlight the importance of phenotype-guided device selection, informed by arrhythmic risk, comorbid burden, and patient-centered goals of care.

## INTRODUCTION

Cardiac resynchronization therapy (CRT) is a cornerstone in the management of heart failure with reduced ejection fraction (HFrEF) and intraventricular conduction delay, particularly in patients with left bundle branch block (LBBB) and a QRS duration ≥130 ms. By restoring electrical synchrony, CRT improves hemodynamic performance, reduces symptoms, lowers hospitalization rates, and improves survival in appropriately selected patients.[1–3]

LBBB occurs in approximately 20% to 30% of patients with HFrEF, and its presence is a major predictor of response to CRT. [4] As the population ages, device-based therapy in older adults is increasingly common, nearly 30% of new CRT implants in the United States are now performed in patients aged ≥75 years. [5] This demographic shift underscores the need to balance potential benefits of CRT against procedural risk and long-term device-related burden in elderly patients.

CRT can be delivered via a pacemaker (CRT-P) or a defibrillator (CRT-D), the latter offering protection against sudden cardiac death through an implantable cardioverter-defibrillator (ICD) component. While CRT-D has demonstrated survival benefit in younger and middle-aged patients,[2,6] it also carries increased cost, procedural complexity, and long-term risks including inappropriate shocks, lead malfunction, and infections. [7,8] These trade-offs may be less favorable in octogenarians, who often face higher competing risk from non-arrhythmic causes of death such as cancer, infection, or progressive heart failure.[9] In this context, the additive value of ICD therapy in elderly CRT recipients remains uncertain.

Despite increasing CRT use in older adults, patients aged ≥80 years have been underrepresented in pivotal CRT and ICD trials. In MADIT-CRT, the median participant age was 64 years; in the RAFT trial, fewer than 5% of patients were older than 75.[10,11] The DANISH trial further questioned the survival benefit of ICDs in patients with nonischemic cardiomyopathy, even in a younger cohort.[12] As a result, guideline recommendations reflect this uncertainty. The 2022 ACC/AHA/HRS guideline acknowledges that “CRT-P may be reasonable in patients with substantial comorbidities or limited life expectancy where defibrillator benefit is uncertain.” [13] Similarly, the 2021 European Society of Cardiology heart failure guidelines emphasize QRS duration and LVEF as primary criteria for CRT candidacy but do not offer device-specific guidance based on age.[14]

Given this evidence gap and clinical ambiguity, real-world data can play a pivotal role in informing device selection in elderly patients. To address this, we conducted a large, multicenter, real-world analysis comparing outcomes between CRT-P and CRT-D in patients aged ≥80 years with HFrEF and LBBB. Using the TriNetX Global Collaborative Network comprising EHR data from 148 health systems, we applied propensity score matching to assess all-cause mortality and hospitalization over a 3-year follow-up period. To our knowledge, this represents one of the largest comparative effectiveness studies of CRT-P versus CRT-D in octogenarians, and offers novel insight into device strategy in a high-risk, under-studied population.

## METHODS

### Data Source and Study Setting

This multicenter, real-world cohort study was conducted using the TriNetX Global Collaborative Network (Cambridge, MA), a federated cloud-based research platform aggregating de-identified electronic health records from 148 health systems, including academic medical centers and large integrated delivery networks across the United States. TriNetX provides access to longitudinal clinical data on demographics, diagnoses, procedures, medications, laboratory values, vital status, and healthcare utilization, updated in real time. All data are de-identified in accordance with the Health Insurance Portability and Accountability Act (HIPAA) Privacy Rule and managed in a secure analytics environment (TriNetX Analytics v6.35).

### Study Design and Population

We identified patients aged ≥80 years with heart failure with reduced ejection fraction (HFrEF; left ventricular ejection fraction [LVEF] ≤35%) and left bundle branch block (LBBB), defined as QRS duration ≥130 milliseconds, who underwent implantation of either a cardiac resynchronization therapy pacemaker (CRT-P) or defibrillator (CRT-D) between January 1, 2010, and April 1, 2024. Only patients with complete demographic and clinical covariates and a minimum of one year of prior EHR activity were included. Those with prior CRT implantation, ambiguous device coding, or incomplete follow-up were excluded.

Device classification was based on procedural codes available in the TriNetX ontology. CRT-P was defined as biventricular pacing without defibrillator capability; CRT-D included biventricular pacing combined with implantable cardioverter-defibrillator (ICD) functionality. The first recorded device implantation was used as the index event, and patients were assigned to the CRT-P or CRT-D group accordingly.

### Propensity Score Matching

A total of 1,240 patients met eligibility criteria (663 CRT-P; 577 CRT-D). To mitigate treatment-selection bias and confounding, we applied 1:1 propensity score matching (PSM) using a logistic regression model with 65 covariates, followed by greedy nearest-neighbor matching without replacement and a caliper width of 0.01 pooled standard deviations. Post-matching, 772 patients were retained for the primary analysis (386 per group). Matching variables included demographics (age, sex, race, ethnicity, geographic region), comorbidities (ischemic cardiomyopathy, atrial fibrillation, chronic kidney disease, diabetes, cancer, stroke), medication use (beta-blockers, ACE inhibitors/ARBs, MRAs, loop diuretics, SGLT2 inhibitors, antiarrhythmics), laboratory data (hemoglobin, creatinine, eGFR, sodium), and healthcare utilization indicators (prior hospitalizations, ED visits, cardiology encounters). Covariate balance was assessed using standardized mean differences (SMDs), with values <0.10 considered indicative of appropriate balance. Full pre- and post-match characteristics and balance diagnostics are presented in Table 1.

**Table 1.**
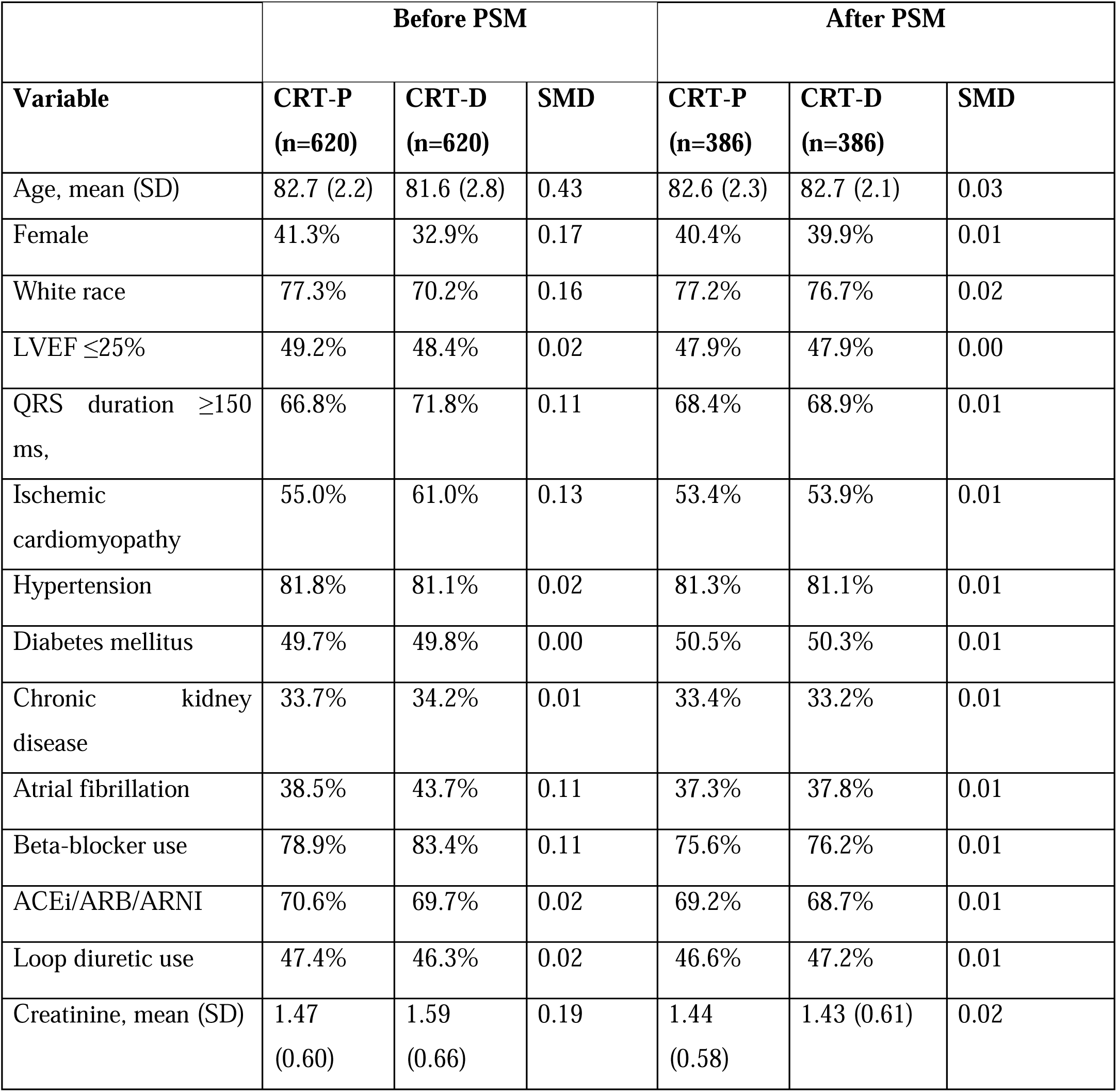
Baseline Characteristics of CRT-P and CRT-D Recipients Before and After Propensity.

In parallel, independent matching was performed for four pre-specified subgroups to permit effect estimation across clinically relevant strata. These included patients with LVEF ≤25% (370 matched patients from 669 eligible), those with LVEF 26–35% (474 matched from 806 eligible), and sex-stratified matched cohorts within CRT-P (N=430, 215 females and 215 males) and CRT-D (N=210, 105 females and 105 males). Each subgroup analysis used the same PSM framework, executed independently to preserve internal balance.

### Outcomes

The primary outcome was all-cause mortality over a three-year follow-up period. Mortality status was ascertained from longitudinal TriNetX records, incorporating EHR-coded death entries across all contributing institutions. Secondary outcomes included the cumulative burden of all-cause hospitalizations per patient over three years. Exploratory analyses also examined time-stratified mortality at 30 days, 90 days, and one year. All follow-up began on the date of CRT implantation and continued until death, last available encounter, or a maximum of three years.

### Statistical Analysis

Categorical variables were compared using chi-square or Fisher’s exact tests, and continuous variables using t-tests or Wilcoxon rank-sum tests as appropriate. Kaplan–Meier survival curves were generated to estimate time-to-event distributions and compared with the log-rank test. Cox proportional hazards models were used to estimate hazard ratios (HRs) with 95% confidence intervals (CIs). Odds ratios (ORs) were calculated for binary mortality endpoints at fixed time points. Hospitalization burden was summarized using means, medians, and interquartile ranges, and compared between groups using t-tests and distributional plots. Statistical significance was defined as p < 0.05 (two-sided). All analyses were conducted using TriNetX Analytics v6.35, a validated environment with standardized epidemiologic and statistical modules.

### Ethics and Data Governance

All data accessed were fully de-identified, and the study was conducted under a data use agreement with TriNetX. Because the study did not involve human subject research as defined by 45 CFR 46.102, Institutional Review Board (IRB) approval was not required. All analyses complied with HIPAA, the TriNetX code of conduct, and international best practices for real-world evidence generation.

## RESULTS

A total of 1,240 octogenarian patients with heart failure with reduced ejection fraction (HFrEF; LVEF ≤35%) and left bundle branch block (LBBB) who received cardiac resynchronization therapy with either a pacemaker (CRT-P) or defibrillator (CRT-D) were initially identified. Following 1:1 propensity score matching (PSM) across clinical and demographic variables, 772 patients (386 CRT-P and 386 CRT-D) were included in the primary analysis (Figure 1, Table 1). Subgroup-specific PSM was conducted separately for ejection fraction (EF ≤25% and 26–35%) and for sex-stratified cohorts within each device type. Sample sizes varied slightly across subgroups due to independent real-time matching algorithms within the TriNetX platform.

**Figure 1.**
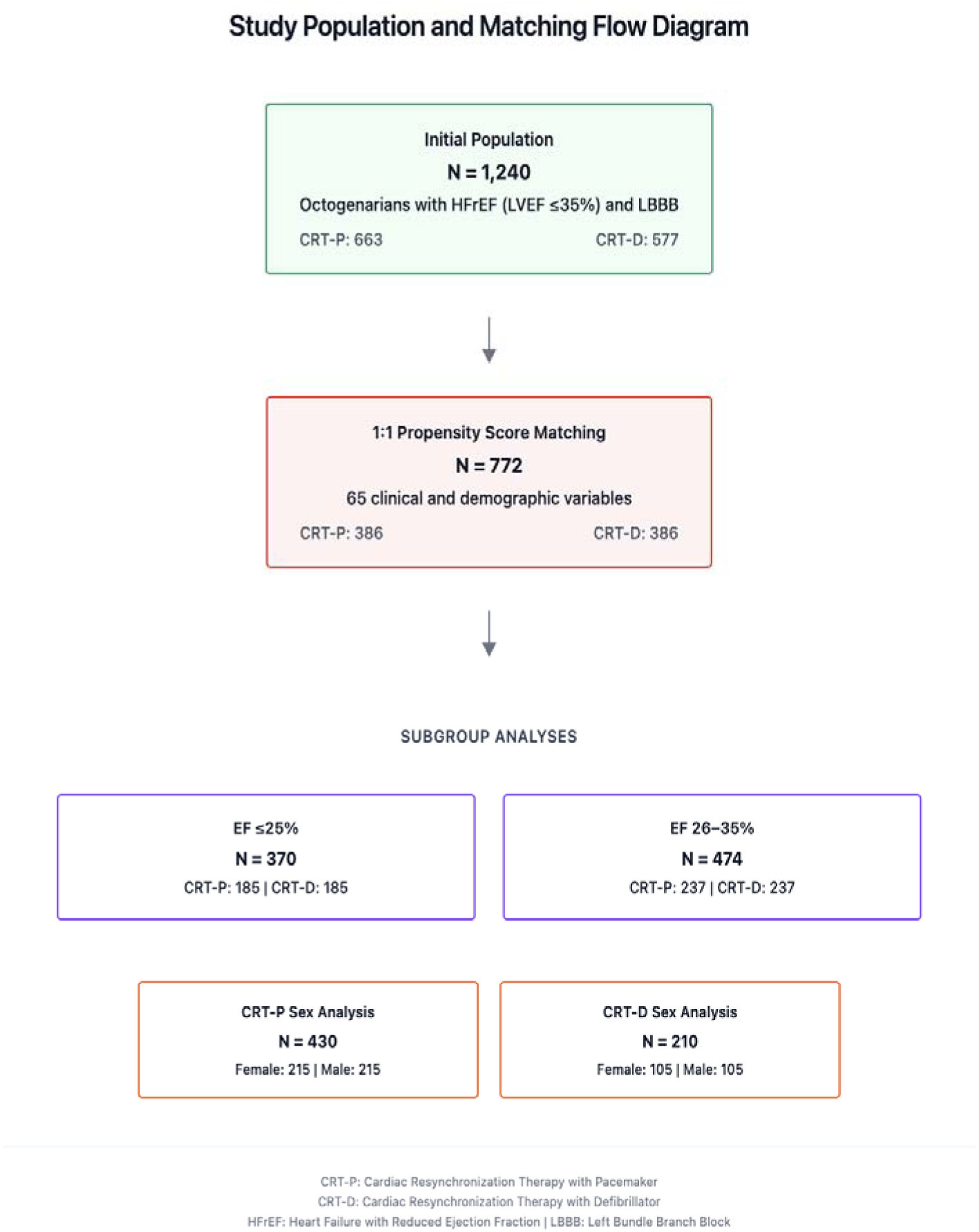
Cohort Architecture and Analytical Structure (Sankey Diagram)

### Primary Outcome: Mortality and Hospitalization Over 3 Years

#### All-Cause Mortality (Matched Cohort, N = 772)

All-cause mortality was similar between CRT-P and CRT-D recipients over a 3-year period. At 30 days, mortality occurred in 8 of 386 CRT-P patients (2.1%) and 7 of 386 CRT-D patients (1.8%) (RR 1.14; OR 1.143, 95% CI 0.418–3.128). At 90 days, 20 deaths (5.2%) were recorded in CRT-P vs 15 deaths (3.9%) in CRT-D (RR 1.31; OR 1.316, 95% CI 0.660–2.625). One-year mortality was 46 of 386 (12.3%) in CRT-P and 39 of 386 (10.1%) in CRT-D (RR 1.22; OR 1.251, 95% CI 0.813–1.926) (Table S1). None of these differences reached statistical significance (all p > 0.05).

At 3 years, 147 deaths occurred in the CRT-P group (147/386, 39.1%) and 141 in CRT-D (141/386, 37.2%), corresponding to an odds ratio of 1.084 (95% CI 0.808–1.454), a hazard ratio of 1.026 (95% CI 0.814–1.293), and a log-rank p-value of 0.829. Median survival was 1,076 days for CRT-P and 1,046 days for CRT-D. Three-year survival probabilities were nearly identical (48.65% vs 48.69%) (Table 2, Figure 2).

**Table 2.**
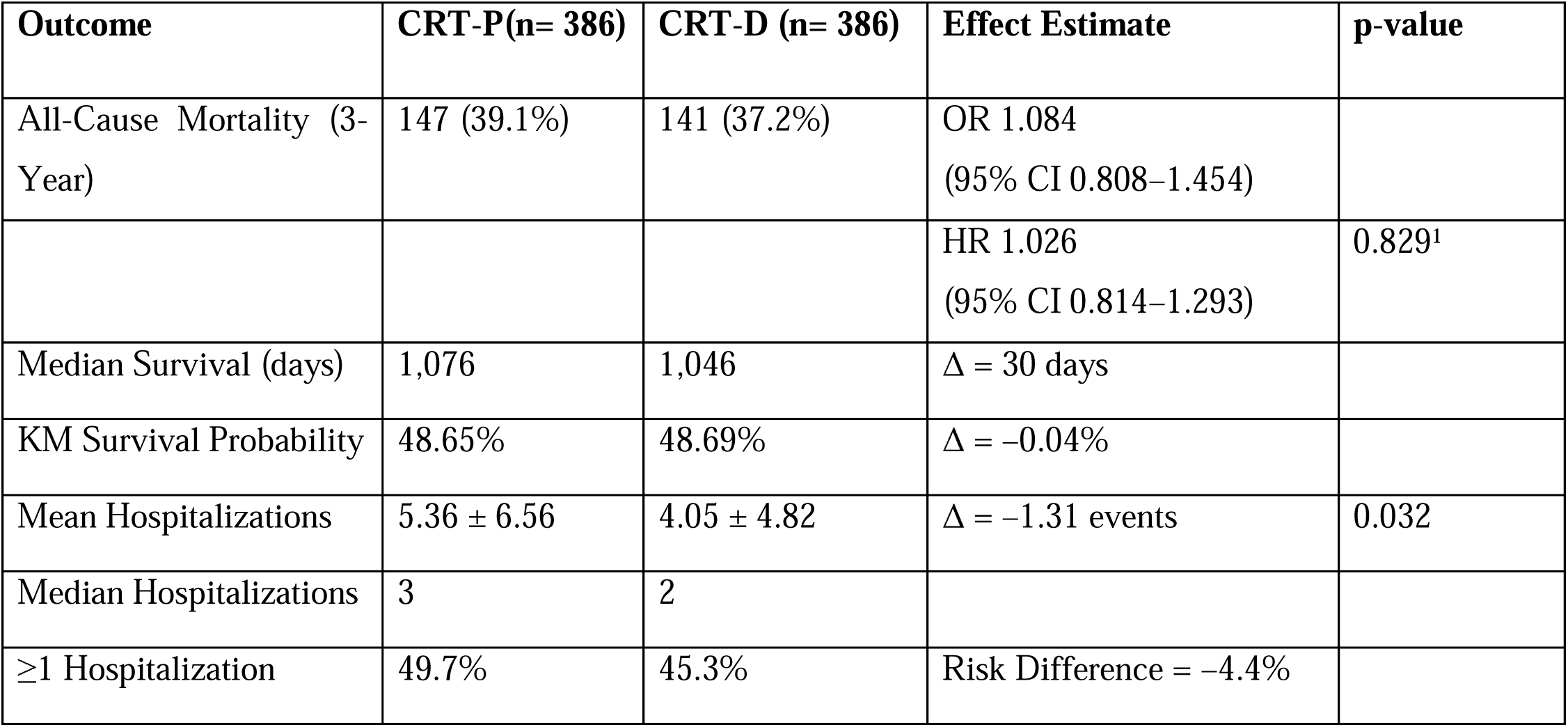
Three-Year All-Cause Mortality and Hospitalization Outcomes in Matched CRT-P and CRT-D Recipients. ^1^ Log-rank p-value from KM survival comparison

**Figure 2.**
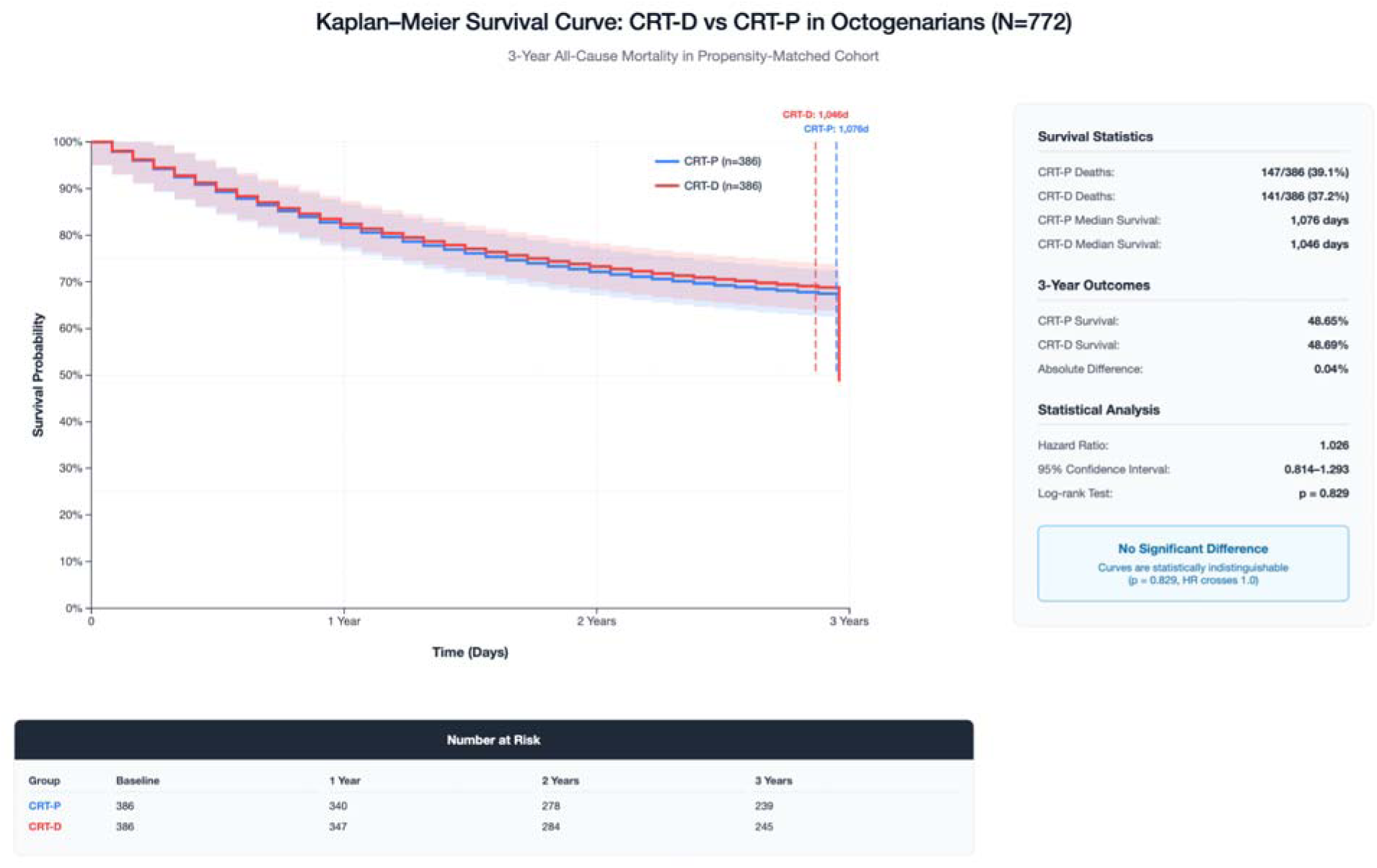
Kaplan-Meier Curve for All-Cause Mortality Over 3 Years: CRT-P vs CRT-D. Kaplan-Meier survival curves with 95% confidence intervals (shaded bands). Log-rank test for comparison. Risk table shows number of patients remaining at each timepoint. HR > 1.0 indicates higher risk for CRT-D vs CRT-P.

#### All-Cause Hospitalization (Matched Cohort, N = 772)

CRT-D recipients experienced significantly fewer hospitalizations over 3 years. The mean number of all-cause hospitalizations per patient was 5.36 (SD 6.56) for CRT-P and 4.05 (SD 4.82) for CRT-D (p = 0.032). Median hospitalizations were 3 in CRT-P and 2 in CRT-D. Additionally, a smaller proportion of CRT-D patients were hospitalized at least once (45.3% vs 49.7%) (Table 2, Figure S3).

### Prespecified Subgroup Analyses: Ejection Fraction Stratification

#### EF ≤25% Cohort (N = 370)

Among patients with EF ≤25%, CRT-D (n = 185) and CRT-P (n = 185) were associated with similar 3-year mortality (60/185 [32.4%] vs 67/185 [36.2%]; risk difference –3.8%). The hazard ratio was 1.113 (95% CI 0.779–1.589; log-rank p = 0.548) (Table S2, Figure S1A).

#### EF 26–35% Cohort (N = 474)

In the EF 26–35% subgroup, 3-year mortality was 77/237 (32.5%) in CRT-D and 82/237 (34.6%) in CRT-P, yielding a risk difference of –2.1% and a hazard ratio of 1.071 (95% CI 0.776–1.479; log-rank p = 0.670) (Table S2, Figure S1B). No statistically significant differences were observed in either EF-defined subgroup.

### Exploratory Analyses: Sex-Stratified Outcomes

#### CRT-P Cohort: Females vs Males (N = 430)

Among CRT-P recipients, 3-year mortality was comparable between females (56/215, 26.0%) and males (59/215, 27.4%). The hazard ratio was 0.914 (95% CI 0.628–1.331; log-rank p = 0.637) (Table S3, Figure S2A).

#### CRT-D Cohort: Females vs Males (N = 210)

Among CRT-D recipients, females had a lower mortality rate than males at 3 years (21/105 [20.0%] vs 31/105 [29.5%]). The hazard ratio was 0.637 (95% CI 0.353–1.149; log-rank p = 0.133) (Table S3, Figure S2B). Although not statistically significant, this trend suggests a potential sex-based differential response to CRT-D that warrants further exploration.

Across all matched cohorts and predefined subgroups, CRT-D was not associated with a statistically significant reduction in all-cause mortality compared to CRT-P. However, CRT-D conferred a significant reduction in cumulative hospitalization burden over a 3-year follow-up. While subgroup analyses of EF and sex did not reveal statistically significant differences, numerical trends favoring CRT-D in patients with lower EF and in female recipients merit further investigation in larger studies (Figure 3, Tables S2–S3).

**Figure 3.**
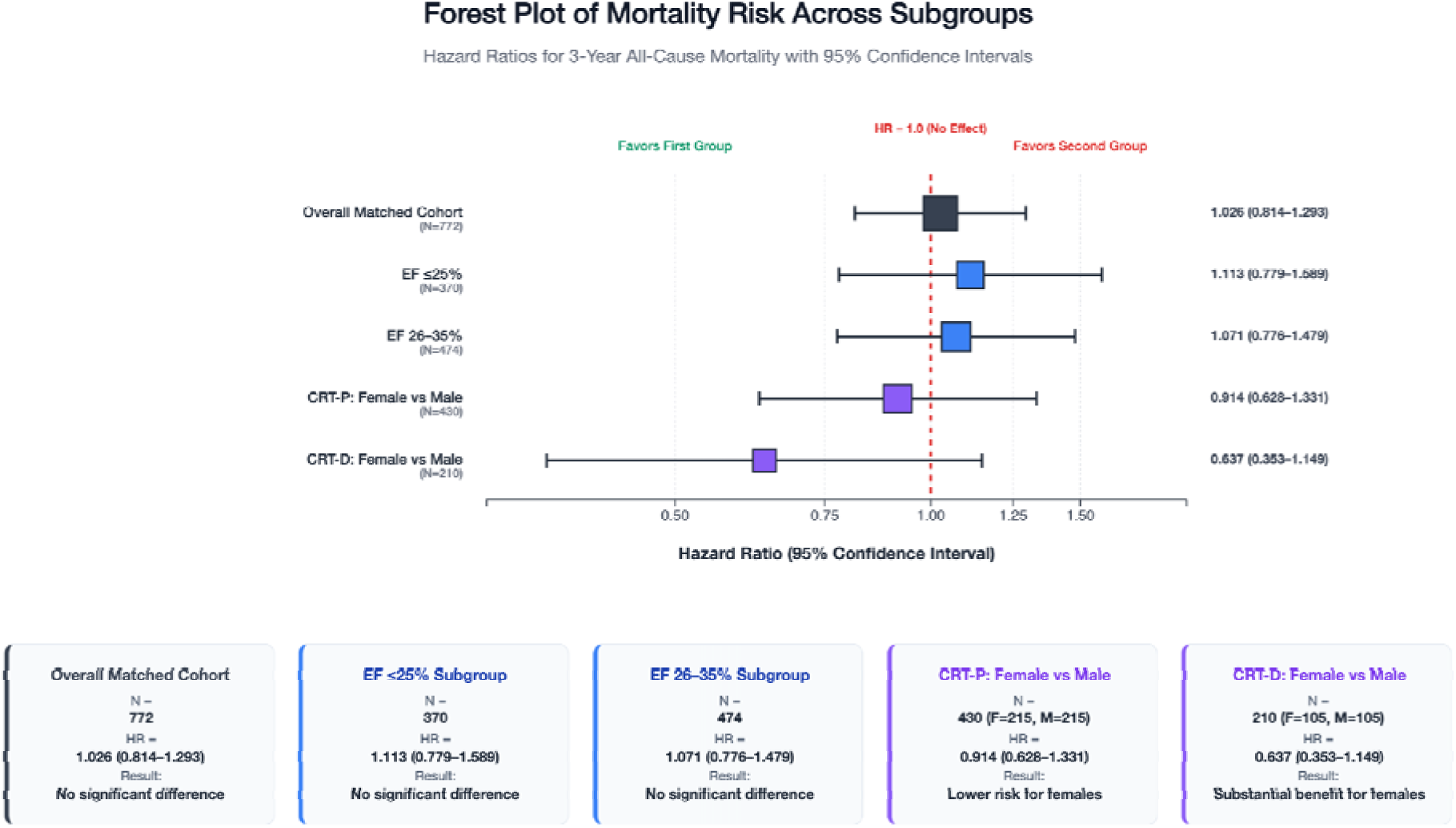
Forest Plot of Hazard Ratios Across All Propensity-Matched Analyses. Clinical Interpretation: All confidence intervals cross HR = 1.0, indicating no statistically significant differences across subgroups. Notable pattern: CRT-D demonstrates greater protective effect in females (HR 0.637) compared to males, suggesting potential sex-based differential response to defibrillator therapy in octogenarians. Forest plot shows hazard ratios with 95% confidence intervals. Square size proportional to sample size. HR < 1.0 favors first group; HR > 1.0 favors second group. Red dashed line indicates null effect (HR = 1.0).

## DISCUSSION

In this large, real-world cohort of octogenarians with heart failure and LVEF ≤35%, we observed no significant difference in all-cause mortality between recipients of CRT-D and CRT-P at 30 days, 90 days, 1 year, or 3 years. This reinforces a growing body of evidence suggesting that the survival advantage of defibrillator therapy attenuates with advancing age, particularly in the presence of substantial comorbidities or competing non-arrhythmic risks. [15–18] While randomized trials established CRT-D efficacy in younger patients with primary prevention indications, observational studies in older adults including those by Yung [19] and Killu [16] have found that non-cardiac mortality may diminish the benefit of defibrillator therapy. These findings support tailoring device selection based on age and clinical phenotype in elderly patients, where CRT-P may offer a more balanced risk-benefit profile.

Although early randomized trials such as MADIT-CRT and COMPANION established the survival benefit of CRT-D in younger populations with broad QRS and reduced EF, [8,20] subgroup analyses and contemporary registry evaluations have noted diminishing returns in elderly patients. [18,21] Our results reinforce these observations, particularly in octogenarians, where competing non-arrhythmic causes of death such as infection, malignancy, or progressive HF may predominate.[18,22] Studies have shown that the incidence of sudden cardiac death declines with age, while non-cardiovascular mortality increases, thus reducing the absolute benefit of ICD-based therapies. [23–26] This aligns with findings from the DANISH trial, which demonstrated no significant mortality benefit of ICD implantation in non-ischemic cardiomyopathy among patients ≥70 years of age. [12]

This study provides novel evidence on differential hospitalization burden between CRT-D and CRT-P in octogenarians, a question seldom addressed in CRT trials. Although all-cause mortality was similar, CRT-D was associated with significantly fewer hospitalizations at 3 years (p = 0.032), a difference not seen at earlier timepoints. This benefit may reflect fewer arrhythmia-related admissions or more proactive care triggered by device alerts and remote monitoring. Remote monitoring has been shown to reduce hospitalizations in CRT-D recipients, [27] though other studies report increased admissions due to inappropriate shocks and device-related complications.[28,29]

One possible explanation is residual selection bias: patients with higher comorbidity or perceived limited survival may have been preferentially assigned to CRT-P, while relatively healthier individuals received CRT-D. Despite robust propensity matching, unmeasured frailty and clinician-driven decisions may still influence these long-term outcomes. These findings underscore the need for individualized CRT strategy in older adults, guided by both physiologic phenotype and goals of care.

The subgroup analysis by EF suggested a 5% absolute reduction in 1-year mortality among patients with EF ≤25% receiving CRT-D, though not statistically significant. This is consistent with mechanistic reasoning and previous CRT trials suggesting more pronounced benefit in those with greater systolic dysfunction. [30,31] In contrast, patients with EF 26–35% had nearly identical outcomes between groups, reinforcing the possibility that CRT-D efficacy may be most relevant in the most severely impaired phenotypes.

In sex-stratified analysis, there was no statistically significant difference in survival between CRT-D and CRT-P recipients within either sex. However, directional trends diverged: among women, 3-year mortality was numerically lower with CRT-D than CRT-P (20.0% vs 26.0%; HR 0.637; p = 0.133), while men showed the reverse (29.5% vs 27.4%; HR 1.09; p = 0.637). Although our study was not powered for interaction testing, this modest absolute difference in female mortality suggests a potential sex-specific response to CRT-D. These findings are consistent with prior meta-analyses and registry studies demonstrating that women with primary prevention ICDs experience fewer appropriate shocks and lower mortality than men, pointing to biologic differences in myocardial substrate, autonomic tone, and arrhythmogenic potential. [32,33]

Current guideline recommendations from the ESC and AHA/ACC/HFSA endorse CRT-D in patients meeting standard criteria for ICD placement and an expected survival of more than one year.[3,34] However, both sets of guidelines emphasize the need for individualized decision-making in older adults with substantial comorbidities or limited life expectancy. Our findings contribute real-world comparative evidence specific to octogenarians with HFrEF and LBBB, suggesting that CRT-P confers comparable survival to CRT-D in this population, with a potentially more favorable risk-benefit profile in appropriately selected patients.

Importantly, CRT-D implantation carries nontrivial risks, particularly in frail or comorbid elderly populations. Studies report inappropriate shock rates of 13–18% at five years,[25] lead malfunction or generator failure in approximately 8–10% of patients,[35] and device-related infections in up to 2%.[36] These complications, especially when occurring in patients with limited arrhythmic risk or reduced physiologic reserve may attenuate the net clinical benefit of CRT-D. Our data underscore the importance of a nuanced, phenotype-informed device selection strategy in older adults that integrates not only LVEF and QRS duration, but also comorbidity burden, frailty indices, and patient-centered care goals.

This study has several important limitations. The TriNetX database lacks adjudicated cause-specific mortality and detailed device-level data, including appropriate and inappropriate ICD therapies. Quality of life measures, frailty indices, and patient-reported functional outcomes were also unavailable. As with all observational analyses, residual confounding and selection bias may persist despite rigorous 1:1 propensity score matching and cohort balancing.

These findings underscore the need for a prospective, randomized trial comparing CRT-D and CRT-P in older adults, particularly those aged ≥80 years. The RESET-CRT trial (NCT03031847) [37] is currently investigating this question. Future research should integrate biologic aging markers, granular comorbidity data, device-specific metrics, and cost-effectiveness modeling to guide precision-based, age-tailored CRT device selection.

## CONCLUSION

In this large, real-world cohort of octogenarians with heart failure and LVEF ≤35%, CRT-D conferred no survival advantage over CRT-P at 30 days, 90 days, 1 year, or 3 years. However, CRT-D was associated with a statistically significant reduction in hospitalization burden at 3 years, a finding that may reflect differences in arrhythmic risk, patient selection, or device response. These results reinforce the need for individualized, phenotype-informed device selection in older adults—balancing arrhythmic risk, comorbid burden, and patient goals of care to guide meaningful therapy.

## Data Availability

All data produced in the present study are available upon reasonable request to the authors

## ABBREVIATIONS

CRT: Cardiac resynchronization therapy
CRT-D: Cardiac resynchronization therapy with defibrillator
CRT-P: Cardiac resynchronization therapy with pacemaker
HFrEF: Heart failure with reduced ejection fraction
LVEF: Left ventricular ejection fraction
LBBB: Left bundle branch block
EF: Ejection fraction
HR: Hazard ratio
CI: Confidence interval
p: p-value

## SUPPLEMENTARY APPENDIX

### Tables

**Table S1.** Time-Dependent All-Cause Mortality: 30-Day, 90-Day, 1-Year, and 3-Year Outcomes.

| **Timepoint** | **CRT-P**<br>**Deaths**<br>**(n=386)** | **CRT-D**<br>**Deaths**<br>**(n=386)** | **Odds Ratio**<br>**(95% CI)** | **Hazard Ratio (95% CI)** | **p-value** |
| --- | --- | --- | --- | --- | --- |
| 30 Days | 8 (2.1%) | 7 (1.8%) | 1.143<br>(0.418–<br>3.128) | — | — |
| 90 Days | 20 (5.2%) | 15 (3.9%) | 1.316<br>(0.660–<br>2.625) | — | — |
| 1 Year | 46 (12.3%) | 39 (10.1%) | 1.251<br>(0.813–<br>1.926) | — | — |
| 3 Years | 147 (39.1%) | 141 (37.2%) | 1.084<br>(0.808–<br>1.454) | 1.026<br>(0.814–<br>1.293) | 0.829 |

**Table S2.** Ejection Fraction–Stratified Mortality Outcomes.

| **EF Subgroup** | **CRT-P Deaths (n)** | **CRT-D Deaths (n)** | **Risk Difference (CRT-D – CRT-P)** | **Hazard Ratio (95% CI)** | **Log-rank p-value** |
| --- | --- | --- | --- | --- | --- |
| ≤25%<br>(N=370) | 67/185<br>(36.2%) | 60/185<br>(32.4%) | –3.8% | 1.113<br>(0.779–1.589) | 0.548 |
| 26–35%<br>(N=474) | 82/237<br>(34.6%) | 77/237<br>(32.5%) | –2.1% | 1.071<br>(0.776–1.479) | 0.670 |

**Table S3.** Sex-Based Mortality Outcomes Within CRT-P and CRT-D Cohorts.

| **Device Type** | **Sex** | **N** | **Deaths (n, %)** | **Hazard Ratio (95% CI)** | **Log-rank p-value** |
| --- | --- | --- | --- | --- | --- |
| CRT-P | Female | 215 | 56 (26.0%) | 0.914<br>(0.628–1.331) | 0.637 |
| CRT-P | Male | 215 | 59 (27.4%) |  |  |
| CRT-D | Female | 105 | 21 (20.0%) | 0.637<br>(0.353–1.149) | 0.133 |
| CRT-D | Male | 105 | 31 (29.5%) |  |  |

### Figures

**Figure S1.**
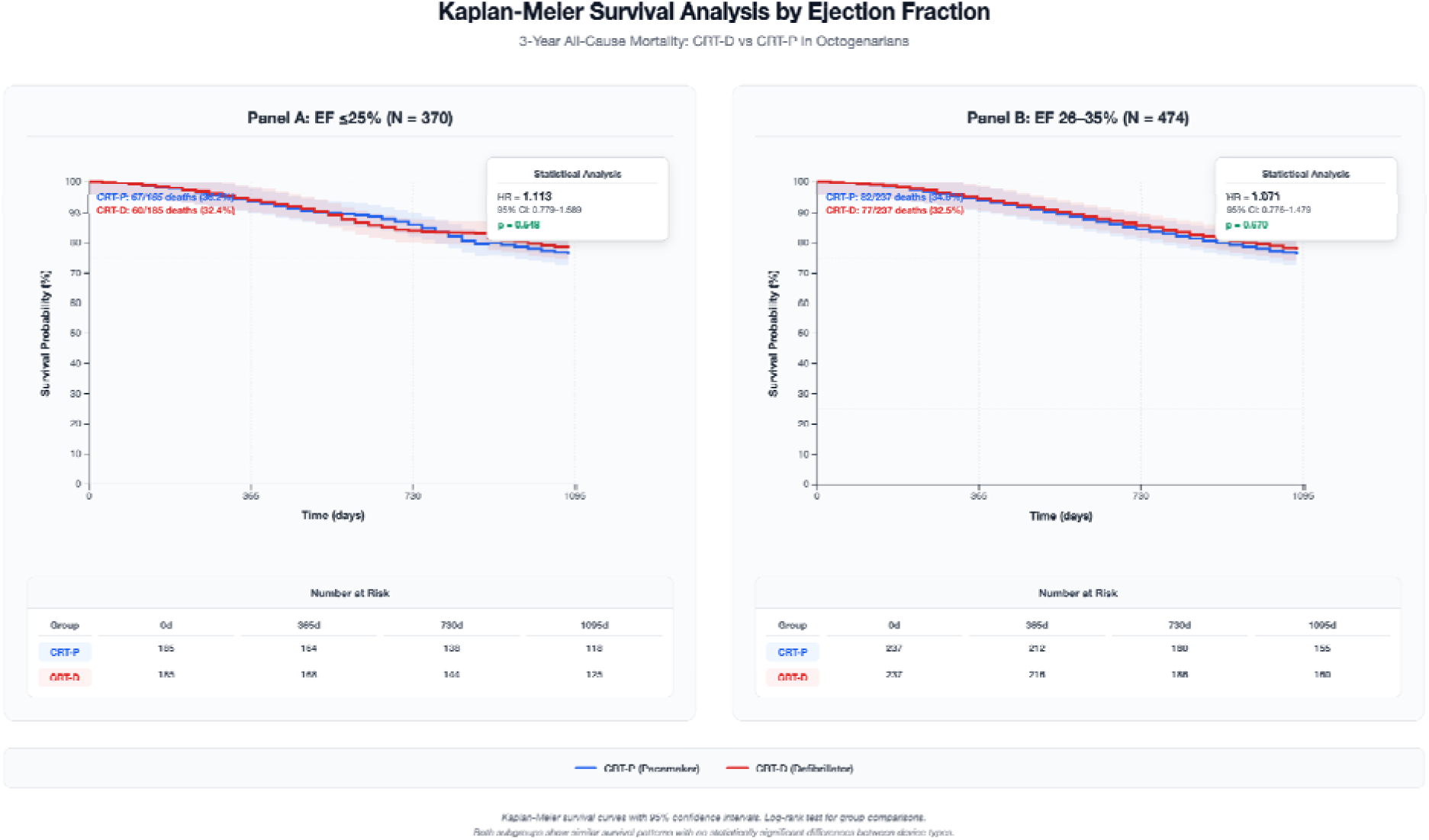
Kaplan–Meier Survival Curves in CRT-D vs CRT-P Recipients Stratified by Ejection Fraction. Kaplan-Meier survival curves with 95% confidence intervals. Log-rank test for group comparisons. Both subgroups show similar survival patterns with no statistically significant differences between device types.

**Figure S2.**
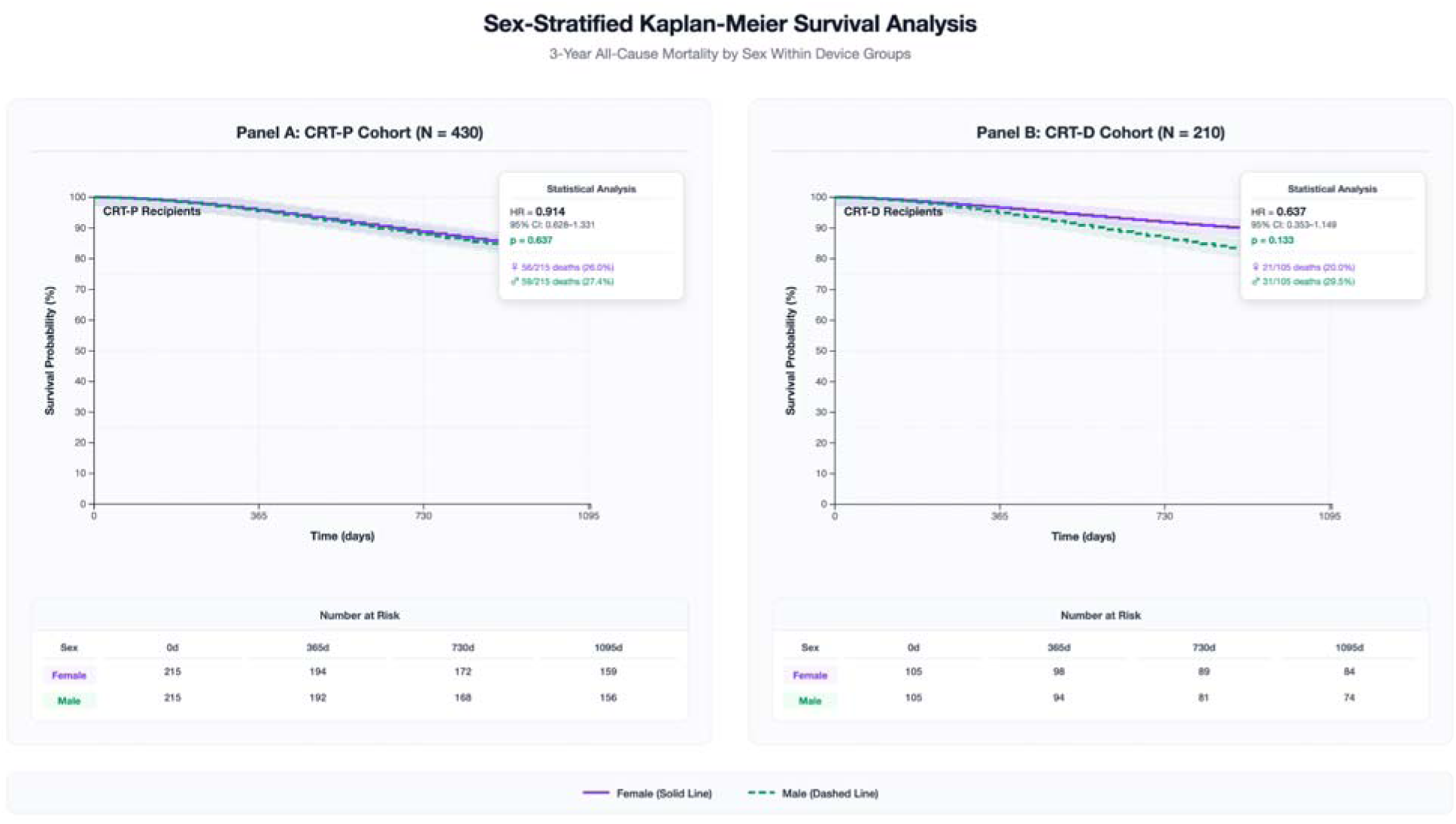
Sex-Based Kaplan–Meier Survival Curves in CRT-P and CRT-D Recipients. Clinical Interpretation: Both device groups show trends favoring female survival, but differences are not statistically significant. Notable pattern: CRT-D demonstrates larger sex-based difference (HR 0.637) compared to CRT-P (HR 0.914), suggesting potential differential benefit of defibrillator therapy in octogenarian females. Kaplan-Meier survival curves comparing female vs male outcomes within each device group. Solid purple lines: females; dashed green lines: males. HR < 1.0 indicates lower risk for females vs males.

**Figure S3.**
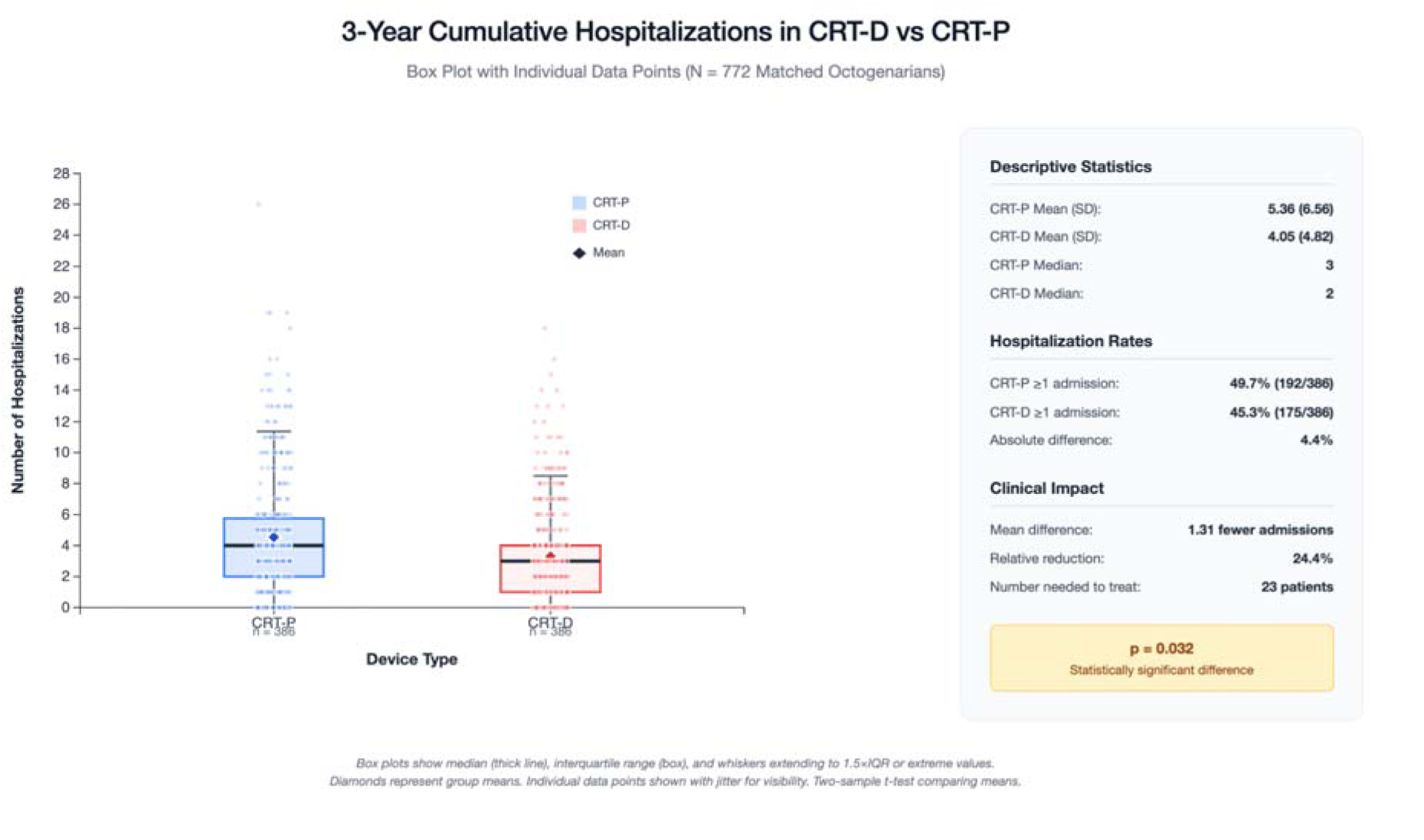
Distribution of All-Cause Hospitalizations by Device Type.

